# The Role of School Environment and Social Interaction in Predicting Student Nurses’ Satisfaction with Academic Programmes: Insights from Two Nursing Training Colleges in Eastern Ghana

**DOI:** 10.64898/2026.04.04.26350149

**Authors:** Michael Narteh Tetteh, Oboshie Anim-Boamah, Atswei Adzo Kwashie

## Abstract

**Background:** Student nurses’ satisfaction with their academic programme is crucial for measuring the success of nursing training institutions. However, in Ghana, studies on student nurses’ satisfaction have mainly focused on clinical learning, neglecting their satisfaction with the academic programme as a whole. This study therefore, assessed the predictors of student nurses’ satisfaction with their academic programme.

**Methods:** A quantitative cross-sectional study design was used in the study. A systematic random sampling technique was employed to recruit 241 student nurses from two Nursing Training Institutions in the Eastern Region of Ghana. The Nursing Student Satisfaction Scale (NSSS) was used for data collection and data was analyzed using Statistical Package for Social Sciences (SPSS) version 27 software.

**Results:** Correlation analysis revealed significant positive associations between satisfaction with curriculum (r = 0.583, p<0.001), faculty role (r = 0.650, p<0.001), social interaction (r=0.680, p<0.001), and overall satisfaction with the academic programme. After adjusting for the school of the student nurses, the school environment (*B=*0.354, p =0.000) and social interaction (*B*=0.291, p=0.001) emerged as significant predictors of student nurses’ satisfaction with their academic programme.

**Conclusion:** The study highlights the need for interventions to enhance the school environment and foster positive social interactions to improve student nurses’ satisfaction with their academic programme.

## 1.0 Introduction

Nursing education plays the role of promoting the growth of the workforce and raising the standards of healthcare and health systems by ensuring that the nurse is provided with all the necessary problem-solving skills (1) to function effectively in an environment characterised by increased workloads, time pressure, and potential stress (2). These problem-solving skills include fundamental knowledge, abilities, and attitudes in the cognitive, emotional and psychomotor domains for scientific problem-solving. Nursing education consists of different academic programmes. An academic programme is a grouping of one or more academic plans around a particular theme, award, or set of admission requirements (3). Some examples include Registered General Nursing (RGN), Registered Midwifery (RM) and Public Health Nursing (PHN) among others.

Student satisfaction with their academic programme is an important metric for assessing the performance of higher educational institutions (4). Since students are at the focal point of every educational institution, their satisfaction with the academic programme serves as a major indicator of institutional progress and also provides feedback for educational administrators to device strategies to continuously improve upon the quality-of-service delivery by the institution. Student satisfaction is a short-term attitude resulting from an evaluation of the students’ educational experience, services and facilities (5). This means that when students perceive that their expectation of educational experiences are met, they may exhibit contentment and portray a positive attitude towards their education thereby actively engaging in all activities of the academic programme. Students’ satisfaction with their academic programme is found to strongly correlate with institutional reputation, student retention and or attrition; hence its assessments have become critical, particularly among non-compulsory higher educational institutions where students are regarded as ‘primary customers’. Further, student nurses’ satisfaction with their academic programme has been found to increase academic performance and retention (6), therefore, understanding how students feel about their academic programme is crucial because dissatisfied students may exhibit signs of distress that can have an impact on their academic and on-the-job performance (7).

Students’ satisfaction with their academic programme has become a crucial element of policy development in most universities across the globe and a prominent focus of researchers in competitive learning environments due to its considerable impact on the success of educational institutes and student enrolment (5). Therefore, higher learning institutions are constantly implementing policies and evolutionary business models to measure performance (8) geared at increasing student satisfaction with higher education quality. Thus, student satisfaction with the academic programme serves as a key element for providing feedback (9).

Factors determining student satisfaction with their academic programme include measurable indicators linked to learning and development outcomes that can be potentially influenced by institutions of higher learning (10) and can be classified into environmental, faculty related issues, social interaction and curriculum. Environmental factors are crucial determinants of satisfaction because adequate infrastructure, access to learning materials, and a conducive learning environment contribute to higher satisfaction levels (11). (12) posit that access to the school building’s emergency exit plan and teaching facilities, (i.e., the computer and clinical laboratories) determine student nurses’ satisfaction with their academic programme. In addition, simulations using contemporary technology and high-fidelity simulations lead to great student satisfaction with their academic programme. Conversely, factors that lead to student nurses’ dissatisfaction with their academic programme include inadequate facilities, insufficient support service, poor lab supplies, and lack of upkeep of the clinical laboratories in response to a large number of students.

Social interactions are the attitudes of the teaching staff and students that allow the curriculum to be successfully implemented. The social interactions that allow the curriculum to be successfully implemented are found to positively predict student satisfaction with their academic programme. Therefore, a democratic leadership style, supportive environment, absence of fear, encouragement of openness and respect for the student as an individual are all elements that contribute to students’ satisfaction with their academic programme. In order to bring about student satisfaction, some components that can be considered for students to navigate their journey include good student-teacher relationship, flexibility and effective communication. Students can gain self-assurance, competence, effective interpersonal communication, and problem-solving abilities in such a setting, which can improve their learning experience. On the contrary, students acquire negative attitudes when they are dissatisfied with their institutions for a variety of reasons, including interpersonal issues, conflict with classmates, and hostile school environments (13). Therefore, (14), argued that interpersonal issues that students face in the classroom make it more difficult for them to articulate the theoretical and practical knowledge that relies on positive interactions between teachers, students, and staff leading to dissatisfaction with their academic programme.

The faculty related issues refer to the teaching staff, particularly the qualifications and preparation of the teachers. Studies have shown that students rate teachers’ expertise as the most influential factor that determines satisfaction and this is determined based on the level of motivation, supervisory relationship with faculty as well as behaviour and efforts of teachers during the duration of the students’ academic programmes. Two factors that likely affect students’ satisfaction with faculty are how helpful the teaching staff is and how well teachers communicate with their classes. Therefore, when faculty are polite, friendly, professional and provide good and prompt feedback, students are actively involved in the teaching and learning process(15). It is obvious that teachers’ teaching styles and methods affect students’ level of satisfaction with the academic programme. For instance, when teachers design teaching and learning strategies that are consistent with students’ abilities and needs as well as the teachers’ own competence, the individual needs of students are met thereby enhancing inclusivity and satisfaction of students (16).

Curriculum is also found to be a significant predictor of student nurses’ satisfaction with their academic programme (17,18) because students are content with their curriculum when they perceive the range of courses it offers to be pertinent to the field of nursing which will help them improve their overall professional growth and analytical skills. A well-sequenced curriculum with subjects of appropriate increasing levels of difficulty positively affects students’ understanding of the content of the curriculum therefore, students are satisfied with their academic programme when these expectations are met. An extensive review identified other student factors such as students’ gender, employment opportunities, learning style, grade point average (GPA), authentic learning, motivation, resilience and financial support as factors that determine students’ satisfaction with their academic programme across tertiary institutions (5,15,19).

In Ghana, however, studies on Nursing education of student nurses focused on aspects of Clinical Learning such as student nurses experiences on clinical learning, challenges of clinical learning education, and teaching and learning clinical competence just to mention a few (20–23) without any research study that investigated student nurses’ satisfaction with their academic programme. The lack of such studies could be a major impediment to the planning and improvement of the academic programme. Therefore, this study is imperative to fill this gap by assessing the predictors of student nurses’ satisfaction with their academic programme in Ghana to enhance programme delivery and academic performance using the Curriculum, Faculty, Social Interaction and Environment (CFSE) model by (24).

## 2.0 MATERIALS AND METHODS

### 2.1 Research Design

Quantitative approach using a cross-sectional study design was used to assess the predictors of student nurses’ satisfaction with their academic programme.

### 2.2 Research Setting

The study was conducted in two (2) public Nursing and Midwifery Training Institutions in the Eastern Region of Ghana that offers Registered General Nursing programme.

### 2.3 Target Population

The target population was student nurses of the two Nursing and Midwifery Training Colleges in the Eastern Region of Ghana.

### 2.4 Inclusion Criteria

All student nurses pursuing General Nursing Programme who had spent at least one year in the selected schools. This group of student nurses had at least a year of experience with the registered general nursing programme in their respective schools and so were in a better position to provide valuable information.

#### Exclusion Criteria

All registered general nursing students who have spent less than a year in the selected Nursing and Midwifery Training Colleges were excluded from the study.

### 2.5 Sample and Sampling Techniques

The number of respondents (sample size) was calculated using the total number of the target population from the two institutions which was four hundred and eighty-five (485) students. Yamane (1967) sample determination formula was used and resulted in a sample size of 219. Further, a 10% of the sample size was calculated and added to cater for non-responses, possible bias and increased participation. Therefore, the sample size was 241 respondents and the number of respondents per school was calculated proportionally by dividing the total number of students in the 2^nd^ and 3^rd^ year classes by the total population (485) and multiplied by the sample size (241). This resulted in a sample of 95 and 146 respondents for the 1^st^ and 2^nd^ selected schools respectively.

A systematic random sampling approach was used in recruiting the respondents. A class list of index numbers of student nurses in the second (2) and third (3) years was obtained with permission from the administration of the two Nursing and Midwifery Training Colleges which resulted in a total of 485 index numbers. This was used as the sampling frame and was divided by the sample size (241) to obtain a k^th^ interval of 2. The first respondent was selected at random between the first index number and the k^th^ index number (interval) thereafter, the researcher selected every 2^nd^ index number (k^th^ interval) on the list (index number on which the k^th^ interval falls) until all the required number of respondents for that class was achieved.

### 2.6 Data Collection Tool and procedure

The instrument selected for the study was the Nursing Student Satisfaction Scale (NSSS) - a validated instrument for measuring students’ satisfaction with their academic programme. The NSSS is divided into four subscales based on the CSFE model: Curriculum nine (9) items; Faculty eight (8) items; Social Interaction, six (6) items; and Learning Environment seven (7) items. Overall satisfaction was measured by one item. Using a six-point Likert scale, the NSSS scores values ranging from 1 (Not satisfied at all) to 6 (Very satisfied). The NSSS had internal consistency scores of 0.96 for the total scale, 0.92 for the curriculum subscale, 0.90 for the faculty subscale, 0.88 for social interaction, and 0.88 for the environment subscale (Chen & Lo, 2015) Data was collected within a month upon receipt of ethical clearance from the Institutional Review Board of the Noguchi Memorial Institute of Medical Research at the University of Ghana, Legon. Participants were given an information sheet, which informed them about the purpose of the study, and they had to sign a consent form to accept their participation in the study. Filled questionnaires were collected from respondents by the close of each day of the data collection.

### 2.7 Validity and Reliability

A data collection instrument that was developed and tested in other studies was adopted for the study. It was pretested using twenty-five (25) registered general nursing students of another Nursing Training School in the Eastern Region. Validity of the instrument was established through face and content validity using the results of the pre-test. Hence, the researcher conducted an extensive literature review and structured the sections of the questionnaire and ensured that all variables under investigation were reviewed. The Cronbach’s alpha was assessed to confirm reliability. Total scale, 0.96 indicated that the adopted questionnaire was reliable.

### 2.8 Data Analysis and Management

The questionnaire responses were screened manually to check their completeness and then coded and uploaded into the Statistical Package for Social Sciences (SPSS) version 27 software. Also, the dataset was saved in the researcher’s personal computer in identifiable folders with security codes for confidentiality. Hard copies of questionnaires were stored in a cabinet under lock and key in the researchers’ office.

Data was analysed using Spearman’s Rho correlation because the variables were ranked or were in an ordinal scale. This determined the correlation between satisfaction with the environment, social interaction, faculty, curriculum and student nurses’ overall satisfaction with their academic programme. Furthermore, hierarchical multiple linear regression was used to assess the extent to which the environment, social interaction, faculty, and curriculum predicted student nurses’ overall satisfaction with their academic programme.

### 2.9 Ethical Consideration

In accordance with the Federal Wide Assurance FWA 00001824 for studies using human respondents, ethical clearance was approved by the Institutional Review Board of the Noguchi Memorial Institute of Medical Research at the University of Ghana, Legon (NMIMR-IRB CPN 032/22-23). An introductory letter was submitted to Health Training Institutions Secretariat, Ghana (HTIS) for institutional clearance and the authorization to select respondents from the schools. Further, consent was sought from each respondent. To ensure privacy and confidentiality, respondents were assigned codes instead of their names to maintain their anonymity. Also, permission was sought from the author of the Nursing Students Satisfaction Scale (NSSS) before the commencement of data collection.

## 3.0 RESULTS

### 3.1 Socio-demographic information of students

Table 3.1 presents an overview of the demographic characteristics of the participants in the study. A total of 241 participants were recruited, 237 student nurses participated in the research while the remaining 4 student nurses did not return their questionnaires. The analysis revealed that the age of the sampled student nurses ranged from 19 to 33 years, with a mean age of 22.47 and a standard deviation of 2.12. Among the participants, the majority (85.70%) were females, while the remaining 14.30% were males. In terms of the academic year, 53.20% of the student nurses were in their second year of nursing studies, while the remaining 46.80% were in their third year. Regarding the educational institutions, 61.60% of the students were studying at the second NMTC, whereas 38.40% were studying at first NMTC.

**Table 3.1:** Socio-demographic information of student nurses (N = 237)

| Variable | Category | Frequency | Percentage | Mean | SD |
| --- | --- | --- | --- | --- | --- |
| Age | 19-33 |  |  | 22.47 | 2.12 |
| Sex | Male | 34 | 14.30 |  |  |
|  | Female | 203 | 85.70 |  |  |
|  | **Total** | **237** | **100.00** |  |  |
| Year of Study | Second year | 126 | 53.20 |  |  |
|  | Third year | 111 | 46.80 |  |  |
|  | **Total** | **237** | **100.00** |  |  |
| School | NMTC 1 | 91 | 38.40 |  |  |
|  | NMTC 2 | 146 | 61.60 |  |  |
|  | **Total** | **237** | **100.00** |  |  |

SD, Standard deviation

### 3.2 Association between study variables

Bivariate analysis using Spearman Rho was conducted to determine the association that exists between the independent variables (curriculum, faculty role, social interaction and school environment) and the dependent variable (satisfaction with the academic programme). Table 3.2 presents the mean, standard deviations and correlation between variables of interest.

**Table 3.2:** Descriptive statistics and correlation between study variables.

|  | **Variable** | **Mean** | **SD** | **1** | **2** | **3** | **4** | **5** |
| --- | --- | --- | --- | --- | --- | --- | --- | --- |
| 1 | Satisfaction with academic programme | 4.658 | 1.401 | — |  |  |  |  |
| 2 | Satisfaction with curriculum | 43.831 | 9.413 | 0.583*** | — |  |  |  |
| 3 | Satisfaction with faculty role | 38.317 | 9.280 | 0.650*** | 0.773*** | — |  |  |
| 4 | Satisfaction with social interaction | 27.165 | 7.703 | 0.680*** | 0.740*** | 0.816*** | — |  |
| 5 | Satisfaction with the school environment | 25.700 | 11.038 | 0.670*** | 0.577*** | 0.682*** | 0.687*** | — |

SD, Standard deviation;
\*\*\* Correlation is significant at the 0.001 level (2-tailed).

The results of the correlation analysis indicated that satisfaction with curriculum has a strong positive and significant association with satisfaction with their academic programme (r = 0.583, p<0.001). This means that as satisfaction with the curriculum increases, satisfaction with the academic programme also increases significantly. Furthermore, student nurses’ satisfaction with faculty roles has a strong positive and significant association with satisfaction with the academic programme (r = 0.650, p<0.001). Thus, as the level of satisfaction of students with the role of faculty increases, there is a corresponding significant increase in their satisfaction with the academic programme. Also, the bivariate analysis shows that satisfaction with social interaction has a strong positive relationship with satisfaction with academic programme (r=0.680, p<0.001). This indicates that as social interaction in schools gets better, students become more satisfied with their academic programme. Finally, satisfaction with the school environment has a strong and significant positive association with the academic programme among student nurses. This gives the signal that improvement in the school environment such as physical structure, library, laboratory, and classroom facilities leads to improved satisfaction with the academic programme among the student nurses.

### 3.3 Predictors of satisfaction with programme

Hierarchical multiple linear regression was conducted to find out the extent to which curriculum, faculty role, social interaction and school environment contribute to explaining student nurses’ satisfaction with their academic programme after controlling for school. In the first step, the school of the respondents was placed in the model (Model 1). In the second step, curriculum, faculty role, social interactions and school environment were placed in the model (Model 2). The results of the regression analysis are presented in Table 3.3. In Model 1, the school of the participants explained 15.3% (Adjusted R square of 0.153) of variance in satisfaction with the academic programme among the student nurses. The addition of curriculum, faculty role, social interaction and school environment to the model (Model 2) explained an additional 39.7% variance (Change in adjusted R square of 0.397) in satisfaction with the academic programme of student nurses. Thus, in Model 2, variables explained 55.5% of the variance in satisfaction with an academic programme which indicates a high predictive power of the regression model. Besides, there was a low mean square error (MSE) of 0.145 which further indicates that the model’s prediction has a relatively low error.

**Table 3.3:** Hierarchical multiple linear regression coefficients of predictors of satisfaction with academic programme.

| Model | Variables | $R^2_{adj}$ | $\Delta R^2_{adj}$ | Unstandardized coefficients | | Standardized coefficients | Significance | Tolerance | VIF |
| --- | --- | --- | --- | --- | --- | --- | --- | --- | --- |
|  |  |  |  | B | Standard error | Beta |  |  |  |
| 1 | Constant | 0.153 | 0.153 | 6.477 | .291 |  | .000 |  |  |
|  | School |  |  | -1.126 | .173 | -.392 | .000 | 1.000 | 1.000 |
| 2 | Constant | 0.551 | 0.397 | .783 | .475 |  | .100 |  |  |
|  | School |  |  | .053 | .154 | .019 | .728 | .680 | 1.471 |
|  | Curriculum |  |  | .012 | .011 | .078 | .286 | .367 | 2.722 |
|  | Faculty role |  |  | .018 | .013 | .119 | .176 | .253 | 3.956 |
|  | Social interaction |  |  | .053 | .015 | .291 | .001 | .275 | 3.636 |
|  | School environment |  |  | .045 | .009 | .354 | .000 | .423 | 2.365 |

$R^2_{adj}$ , Adjusted R square; $\Delta R^2_{adj}$ , R square change;
B, Unstandardised Coefficient Beta; VIF, Variance Inflation Factor;
Dependable variable: Satisfaction with the academic programme

The assumptions of regression were scrutinised utilising the recommendations of (25) as well as Williams, Grajales, and Kurkiewicz (2019). Partial and scatter plots of the predictors within the model were employed to confirm that linearity and homoscedasticity assumptions were not breached. Furthermore, tolerance values exceeding 0.1 and Variance Inflation Factors (VIF) values less than 10 were discovered, thereby indicating that issues of multicollinearity do not exist within the regression model(26). Additionally, with a Durbin-Watson statistic of 1.947, it can be inferred that there are no issues of autocorrelation in the data. The violation of the normality of the residual distribution assumption (Kolmogorov-Smirnov = 0.242, p = 0.000, Shapiro-Wilk = 0.834, p = 0.000) was detected. The violation of this assumption warrants caution when interpreting the results of the regression analysis. However, as posited by William et al. (2019), compliance with the assumption of normally distributed errors is not required to obtain a reliable and unbiased coefficient of regression, if other regression assumptions are met. Moreover, in a sample greater than 200, the central theorem guarantees that residual distribution approaches a normal distribution (Osborne & Waters, 2019; Williams et al., 2019). Consequently, regression models are relatively robust to the normal residual distribution assumption (Osborne & Waters, 2019). The regression results indicate that when adjusting for the school of the student nurses, social interaction (*B*=0.291, p=0.001) and school environment (*B=*0.354, p =0.000) make a significant contribution to explaining student nurses’ satisfaction with their academic programme. Also, the school environment is the highest predictor of student nurses’ satisfaction with the academic programme.

## 4.0 DISCUSSION

The purpose of this study was to assess the predictors of student nurses’ satisfaction with their academic programme in the Eastern region of Ghana. The study found that satisfaction with the school environment had a strong and significant positive association with nursing students’ satisfaction with their academic programme (r=0.67, p<0.001). This gives the signal that improvement in the school environment such as physical structure, library, laboratory, and classroom facilities could lead to improved satisfaction with the academic programme among the student nurses. According to (27), availability of appropriate infrastructure, support facilities, equipment procurement, and maintenance services increases student nurses’ satisfaction with their academic programme and vice versa. This could be because the availability of adequate resources in the school environment ensured that all the necessary items needed to study are available to students, hence, increasing their level of satisfaction with their academic programme.

Also, in the current findings, low satisfaction of students with their academic programme may be due to the poor setup of their academic environment and the unavailability of some facilities in their school environment. The current findings imply that student nurses’ level of satisfaction with their academic programme could decline if there is inadequate infrastructure, learning materials and a poor learning environment (28). Also, there could be a decline in their academic performance, which could lead to production of less motivated and competent staff nurses and poor patient care (29). Besides, a low level of satisfaction with the academic programme among student nurses due to a poor academic environment could contribute to increased levels of stress and burnout among students impacting their mental and emotional well-being (30). Further, after adjusting for the school of the student nurses, the school environment was the highest predictor of student nurses’ satisfaction with their academic programme as it explained 35.4% of the variance in student nurses’ overall satisfaction with their academic programme (*B=*0.354, p =0.000) compared to social interaction (*B*=0.291, p=0.001). This finding implies that the nursing school environment has a higher magnitude of impact on students’ chances of becoming satisfied with their academic programme. Thus, any attempt to undermine the provision and maintenance of adequate resources and conducive teaching and learning environment in nursing schools could affect their satisfaction level of the academic programme.

The study also found that student nurses’ satisfaction with faculty roles has a strong positive and significant association with satisfaction with the academic programme (r=0.65, p<0.001). Thus, as the level of satisfaction of students with the role of faculty increases, there is a corresponding significant increase in their satisfaction with the academic programme. Moreover, poor satisfaction with the school faculty could lead to low satisfaction with the academic programme. The possible reason for this relationship could be that, because faculty members are responsible for delivering lectures, organising clinical experiences, and guiding students in their learning process, when students are satisfied with the competence and effectiveness of their faculty members, they are more likely to feel engaged, motivated, and supported in their academic journey. This positive learning experience can contribute to overall satisfaction with the academic programme. Moreover, faculty members usually serve as role models for student nurses, hence, when students perceive their faculty as knowledgeable, experienced, and passionate about nursing, it could inspire and motivate them. Also, positive role modelling by faculty could enhance students’ confidence in their chosen career path and increase their satisfaction with the academic programme (31,32). Similarly, (33) found that students who had high levels of satisfaction with their faculty were highly satisfied with their academic programme. Likewise, a study by (34), which found excellent student satisfaction with their academic programme because their lecturers promptly provided feedback on assignments. On the contrary, (35) found low student nurses’ satisfaction with their faculty which negatively affected nursing students’ satisfaction with the academic programme. This was due to poor mentoring of students by school staff.

In the regression analysis, satisfaction with the faculty role did not make a significant contribution to explaining student nurses’ overall satisfaction with their academic programme (*B=*0.119, p =0.176). This suggests that other factors may have a stronger influence on their satisfaction levels. Several reasons may explain this result. Firstly, students’ satisfaction may be influenced more by the broader school environment, such as facilities, resources, and the overall learning atmosphere, rather than solely by their satisfaction with individual faculty members. While faculty members play a crucial role in education, their impact may be one piece of the larger puzzle that shapes overall satisfaction with the academic programme.

Furthermore, the study found that satisfaction with social interaction has a strong positive relationship with satisfaction with the academic programme (r=0.680, p<0.001). This indicates that as social interaction in schools gets better, students become more satisfied with their academic programme. The possible reason is that, perhaps students who have a good relationship with their tutors may develop high confidence that enables them to ask questions for effective learning. Studies show that a strong relationship between a tutor and a student promotes effective communication that enables tutors to understand students’ unique needs, learning styles, and goals (36–38). This helps tutors tailor their teaching methods accordingly. This understanding creates a conducive learning environment and enhances student engagement and satisfaction (39). Also, when students feel comfortable with their tutors, they are more likely to seek help, ask questions, and share their concerns (40). Besides, tutors who provide support and guidance contribute to students’ confidence and overall satisfaction with their academic programme (41). A good tutor-student relationship offers emotional support, creating a safe space for students to express their concerns, frustrations, and anxieties. Tutors who provide empathy, encouragement, and guidance help students navigate challenges, develop resilience, and maintain their satisfaction with the academic programme (42). Similarly, (33) recorded high satisfaction with the academic programme among their participants due to their participants’ high level of satisfaction with their relationship with tutors. It was revealed in their study that their participants reported that their tutors were approachable and made an attempt to listen to them. Likewise, (34) found high satisfaction with the academic programme due to student satisfaction with lecturer-students’ relationships. On the contrary, (35) observed low satisfaction with the academic programme due to poor relationship between tutors and students. In this study, satisfaction with academic programme increases when there is a good tutor-student relationship and declines with a poor tutor-student relationship. After adjusting for confounding variables, social interaction made a significant contribution to explaining student nurses’ satisfaction with their academic programme. This means that, tutor-student relationship plays a critical role in ensuring student nurses’ satisfaction with their academic programme. Hence, any factor that compromises a good relationship between tutors and student nurses could affect their level of satisfaction with their academic programme.

Social interaction plays a significant role in explaining student nurses’ satisfaction with academic programmes and this can be attributed to several factors. Positive and engaging interactions between tutors and students create a supportive and conducive learning environment (36). From this study, when tutors actively interact with students, provide guidance, and offer personalized support, it enhances students’ understanding, motivation, and overall satisfaction with the academic programme. In addition, social interaction promotes effective communication and feedback (43). Regular and meaningful communication between the tutors and student nurses allowed for clarifying doubts, discussing challenging topics, and receiving constructive feedback as expressed by (36). Thereby fostering a sense of belonging, encouraging active participation, and contributing to students’ perceived value of the academic programme. Focusing on improving social interaction can lead to an enhanced learning experience for student nurses. Thus, by fostering a supportive and interactive environment, students are more likely to engage actively in their studies, leading to improved understanding, knowledge retention, and overall academic achievement (38).

From the study, satisfaction with curriculum had a strong positive and significant association with satisfaction with the academic programme. As satisfaction with the curriculum increases, satisfaction with the academic programme also increases significantly. Thus, in instances where student nurses perceive their curriculum to be effective in preparing them for their careers, they are more likely to feel satisfied with their academic programme and vice versa. Perhaps, when student nurses feel their curriculum is good enough to prepare them for their career goals, they may commit to their academic activities, hence, increasing their level of satisfaction with their academic programme(44). Similarly, (24)Chen and Lo (2015) revealed that student nurses in the US were satisfied with their academic programme because the participants were happy with the curriculum as they felt it was pertinent to the field of nursing. (33) found that students were happier with their curriculum because they thought the range of courses it offered would help them improve their overall professional growth and analytical skills. These align with the current study indicating that if students perceive the curriculum as well-designed, relevant, comprehensive and effective in meeting their learning needs, they will be more satisfied with their academic programme. A satisfying curriculum can enhance students’ engagement and motivation, leading to better learning outcomes. Satisfied students are more likely to actively participate in classroom activities, clinical practice, and self-directed learning, which can improve their knowledge, skills and their overall level of satisfaction with their academic programme. However, although the regression analysis revealed that satisfaction with curriculum did not make a significant contribution to explaining student nurses’ overall satisfaction with their academic programme, other factors may have a stronger influence on student nurses’ overall satisfaction levels. This can be explained by the fact that, while the curriculum plays a pivotal role in guiding students’ educational journey, their overall satisfaction may be influenced by a combination of various factors beyond the curriculum itself. The broader school environment, teaching methods, resources, and support services may have a more significant impact on students’ satisfaction with their academic programme. It is therefore imperative to consider a holistic approach to enhancing student nurses’ overall satisfaction with their academic programme by providing adequate and modern infrastructure, fostering social interaction among faculty and students, reviewing the curriculum regularly to reflect current trends in nursing practice and providing opportunities for continuous professional development for faculty on instructional methodology. Much as the curriculum is a fundamental component of nursing education, it is essential to recognize and address other factors that contribute to student satisfaction. Educational institutions should focus on creating a positive and supportive school environment, ensuring the availability of adequate resources, fostering effective teaching methods, and providing robust student support services. With a comprehensive approach, institutions can create an environment that supports student nurses’ educational journey, meets their needs and expectations, and ultimately enhances their overall satisfaction with the academic programme. Collaborative efforts among faculty, administrators, and students are crucial in ensuring that the academic programme aligns with students’ goals and aspirations, leading to a more fulfilling educational experience.

## 5.0 CONCLUSION

The findings demonstrate that satisfaction with the school environment, curriculum, faculty and social interactions wield a potent and substantial positive association with student nurses’ satisfaction with their academic programme. Both the school environment and social interaction are the significant and formidable contributors to explaining student nurses’ satisfaction with their academic programme. It is therefore, recommended that nursing training institutions be provided with sufficient funding to acquire the necessary resources and infrastructure to provide a high-quality educational experience. There should also be periodic curricular review to reflect current best practices and incorporate emerging trends in healthcare. Further, Nursing Training Colleges should promote positive interactions between staff and students through group activities, mentorship programmes, and peer support networks to create a sense of belonging and camaraderie among students.

## Data Availability

All relevant data are within the manuscript and its supporting information files.

## Declarations

### Declaration of Conflicts of Interest

The authors declare that there is no conflict of interest.

### Funding

This research received no funding from any funding agency in the public, commercial, or not-for-profit sectors. This is a publication from an MPhil Thesis.

### Consent for Publication

The data was collected from student nurses who gave their informed consent for the study. The ethical approval certificate is (NMIMR-IRB CPN 032/22-23) All authors agree to the publication.

### Ethical Approval

The study gained institutional ethics approval from the Institutional Review Board of Noguchi Memorial Institute for Medical Research, University of Ghana, Legon (NMIMR-IRB CPN 032/22-23). Participation was voluntary and all participants provided their free informed consent. All ethical protocols were observed.

### Availability of Data and Materials

Data is provided within the manuscript and in supplementary information files. The data used in this manuscript were part of a MPhil Thesis research and can be made available from the corresponding author upon request.

### Competing Interests

The authors declare that, there are no competing interests in this article.

### Funding

No funding was received in this work. The publication is from research conducted as part of the requirement for the award of MPhil degree in Nursing.

### Clinical trial number

Not Applicable.

### Author Contributions

Michael Tetteh conducted data collection, and analysis, and drafted the manuscript. Dr. Anim-Boamah and Dr. Kwashie contributed to the writing of the discussion, edited, and reviewed the article for intellectual content. All authors approved the final version of the article.

## Acknowledgement

The authors express their gratitude to Dr. Charles Teye and Mr. Charles Djorbua for supporting in the coding and data entry into the SPSS. We are grateful to all the participants from the Schools. The research supervisory team is duly acknowledged.

## Authors Biodata

**Michael Narteh Tetteh MPHIL, PGDE, BSC, RN**

^1^Nursing and Midwifery Training College, Atibie-Kwahu, Phone Number: +233209574864

P. O. Box 253, Atibie, Kwahu.

**Oboshie Anim-Boamah PhD, MPHIL, MED, PGDE, RN, FWACN**

School of Nursing and Midwifery, University of Ghana, Phone Number: +233208232127

P. O. Box LG 43, Legon, Accra.

**Atswei Adzo Kwashie PhD, MPHIL, SRN- corresponding author**

School of Nursing and Midwifery, University of Ghana Phone Number: +233244276317

P. O. Box LG 43, Legon, Accra.

## Notes

### Competing Interest Statement

The authors have declared no competing interest.

### Funding Statement

The author(s) received no specific funding for this work.

### Author Declarations

In accordance with the Federal Wide Assurance FWA 00001824 for studies using human respondents, ethical clearance was approved by the Institutional Review Board of the Noguchi Memorial Institute of Medical Research at the University of Ghana, Legon (NMIMR-IRB CPN 032/22-23)

